# Opioid Overdose Death Prediction with Graph Neural Networks

**DOI:** 10.64898/2026.03.18.26348454

**Authors:** Xianhui Chen, Zishan Gu, John Myers, Joanne Kim, Changchang Yin, Naleef Fareed, Neena Thomas, Soledad Fernández, Ping Zhang

**Affiliations:** Computer Science and Engineering, The Ohio State University, Columbus, Ohio, USA; Department of Biomedical Informatics, The Ohio State University, Columbus, Ohio, USA; Center for Biostatistics, The Ohio State University, Columbus, Ohio, USA

## Abstract

The opioid crisis has severely impacted Ohio, with overdose death rates surpassing national averages and disproportionately affecting rural and Appalachian regions. Accurately predicting county-level opioid overdose (OD) deaths is critical for timely intervention but remains challenging due to the wide differences in opioid OD deaths between large and small counties. We propose a Spatial-Temporal Graph Neural Network (ST-GNN) framework that integrates graph neural networks (GNNs) to capture spatial relationships between counties and Long Short-Term Memory (LSTM) networks to model temporal dynamics. Using quarterly OD death data from Q1 2017 to Q2 2023 for 88 Ohio counties, we incorporate a nine-dimensional dynamic feature set, including naloxone administration events and high-risk opioid prescribing, along with a static Social Determinants of Health (SDoH) index. Compared to traditional statistical models and temporal deep learning baselines, our ST-GNN demonstrates superior performance, particularly in larger counties, while a classification-based strategy improves predictions for small counties, leading to more stable and reliable results. Our findings emphasize the need for spatial-temporal modeling and customized training to enhance public health decision-making in addressing the opioid crisis.

## 1 Introduction

The opioid crisis in Ohio has reached critical levels, making it one of the hardest-hit states in the United States [1, 2]. In 2020 alone, Ohio reported 5,204 opioid overdose (OD) deaths, translating to a rate of 42.7 deaths per 100,000 individuals—more than 1.5 times the national average [3, 4]. As reported by Ohio unintentional drug overdose report [5, 6], this crisis is exacerbated by the widespread availability of synthetic opioids, such as fentanyl, which accounted for 81% of the state’s OD deaths in 2020. The crisis disproportionately impacts rural Appalachian counties and urban areas, indicating that it affects everyone [7].

Timely resource allocation and response are essential for preventing the further escalation of overdose deaths. Consequently, many studies focus on developing predictive models that enable timely interventions [8, 9, 10]. Although these methods have demonstrated promising performance, there are three limitations: *(1) They do not integrate the spatial relationships inherent in geographic information*. Given the natural graph structure of geographic data, Graph Neural Networks (GNNs) have been effectively employed to address this limitation in predicting the spread of pandemics and other dynamic phenomena [11, 12, 13]. However, their application in predicting opioid overdose deaths requires more attention. More specifically, limited efforts have been made to capture the interplay of temporal and spatial factors at a granular level, which is critical for understanding and predicting overdose death rates. *(2) Furthermore, existing methods fail to integrate static information with dynamic data*. While research [14, 15] emphasizes the role of social determinants of health (SDoH), such as economic stability and access to healthcare, there is a lack of methods that jointly integrate these static SDoH indicators with dynamic time-series data and spatial geographic relationships. *(3) Current studies [16, 17, 18] apply the same loss function to both large and small counties, leading to unfair modeling*. The opioid overdose death rates in large and small counties differ widely due to disparities in population size. A general loss function prevents the model from learning distinct patterns for different county sizes, resulting in considerable fluctuations, even for the same county at different prediction time points.

To address these challenges, we propose the Spatial-Temporal Graph Neural Network (ST-GNN) framework, a novel approach for predicting opioid overdose death rates at both county and regional levels. Notably, the data used in our study has a six-month lag due to the delay in data caused by the government’s processing time [19]. Therefore, our goal is to predict opioid overdose deaths with a six-month gap and provide real-time predictions. Our framework leverages the strengths of GNNs to model spatial relationships between counties, augmented by Long Short-Term Memory (LSTM) networks to capture temporal dynamics. ST-GNN incorporates adjacency graphs to encode spatial dependencies and jointly processes static SDoH features alongside dynamic temporal data. Additionally, we propose a weighted joint training loss that combines two sub-loss functions for large counties and small counties. In particular, this study uses Ohio’s quarterly opioid overdose data from 2017 to 2023, enriched with dynamic features such as EMS naloxone administration events and static SDoH features, to train and evaluate the ST-GNN framework. By jointly training these components, the ST-GNN framework dynamically models the evolution of opioid overdose deaths across time and geography. This method effectively combines static and dynamic features, enabling robust and actionable predictions. Furthermore, to account for disparities among counties with varying population sizes, we tailor prediction tasks accordingly with a joint training loss: for small counties where monthly or quarterly death counts are close to three, we formulate a binary classification task to predict whether the death count will exceed three. In contrast, for larger counties, we perform a standard regression task to predict the actual death counts. Our work not only advances the state-of-the-art in predictive modeling for opioid overdose deaths but also provides a scalable and flexible solution for addressing public health crises driven by complex spatial-temporal phenomena.

Experiments conducted on data reported by the Ohio Department of Health (ODH) demonstrate that our proposed method outperforms all baseline models for both large and small counties. For large counties, our method achieves an RMSE of 9.149 and an SMAPE of 0.242. For small counties, it achieves an ROC-AUC of 0.738 and an F1 score of 0.579.

Overall, our contributions can be summarized as follows:

- We developed a GNN-based framework that jointly captures temporal and spatial dependencies for predicting county-level opioid overdose death rates.
- We proposed a weighted joint training loss to perform predictive tasks on counties with different population sizes.
- We leveraged comprehensive data from ODH, including dynamic opioid-related metrics and static SDoH indices, to validate model performance. Our framework achieves the best performance with GAT being the GNN model compared with both traditional statistical methods and state-of-the-art spatio-temporal methods.

## 2. Preliminary Studies

Here, we present preliminary studies aimed at investigating the challenges of predicting county-level opioid overdose deaths using deep learning models. Specifically, we employ Long Short-Term Memory (LSTM) [20], a widely used model for time-series forecasting, using only the death count as input. The objective is to observe how deep learning methods perform in this task when provided with limited temporal data and variations across counties. Our experiments are designed to address the following key questions: Is the data from a single county sufficient to train an effective deep learning model? Can training be enhanced by augmenting a county’s data with data points from other counties? Do LSTM models exhibit performance disparities across different counties? If so, what are the underlying factors contributing to this performance gap?

### 2.1 Data Augmentation

We conducted several experiments and summarized the results in Table 1. Each row of the table presents the performance metrics—Root Mean Squared Error (RMSE) and Mean Absolute Percentage Error (MAPE)—for LSTM models trained using different subsets of data. We evaluate models trained with data from single counties, first-order neighbor counties, counties grouped by similar types, and all counties. The following paragraphs will discuss these settings and their corresponding results in detail.

**Table 1.**
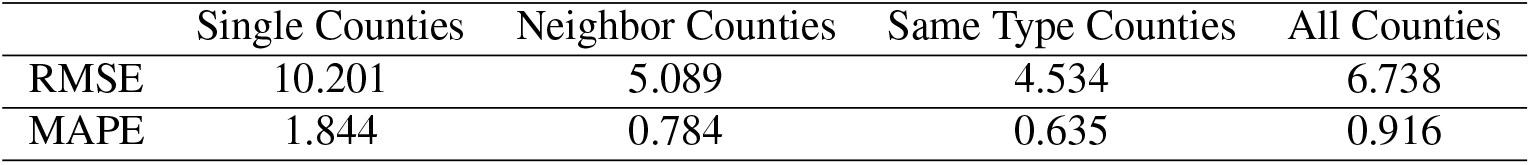
LSTM models trained with different sets of training data.

|  | Single Counties | Neighbor Counties | Same Type Counties | All Counties |
| --- | --- | --- | --- | --- |
| RMSE | 10.201 | 5.089 | 4.534 | 6.738 |
| MAPE | 1.844 | 0.784 | 0.635 | 0.916 |

We first trained individual LSTM models for each of the 88 counties, resulting in 88 separate models. As shown in the table, this configuration produced the worst overall performance, with the highest RMSE and MAPE values. The poor performance can be attributed to overfitting, as the limited amount of training data from a single county lacks sufficient diversity to generalize well. The model effectively memorizes patterns specific to the county but struggles when faced with unseen data, leading to suboptimal predictions. To address the overfitting observed with single-county models, we next trained a single LSTM model using data aggregated from all 88 counties. This approach yielded improved performance, with a noticeable reduction in both RMSE and MAPE. The improvement highlights the benefit of data augmentation, where combining data from multiple counties introduces greater diversity, thus reducing overfitting and enhancing the model’s generalization capability. However, while this method shows improvement, its gains are still limited, suggesting that naively pooling data across all counties without distinguishing them is not optimal.

We then explored whether leveraging spatial relationships could further improve model performance by training individual models for each county using data from both itself and its neighboring counties. This approach, similar to the single-county setup, resulted in 88 models, but with improved performance relative to the single-county models. By including geographically relevant data, the models were better equipped to learn shared temporal patterns, which alleviated overfitting and yielded more accurate predictions. This suggests that incorporating spatially correlated information can be beneficial for enhancing prediction accuracy. To further refine the data augmentation strategy, we categorized the counties into four types: Appalachian, rural, suburban, and urban, and trained four separate LSTM models—one for each type. This grouping method also provided performance gains, as shown by reductions in RMSE and MAPE. By grouping counties with similar characteristics, the models were better able to generalize within each category and capture region-specific temporal patterns, leading to enhanced predictive performance.

#### Key Observations

Overall, the results indicate that training models using data from a single county is insufficient due to overfitting and lack of generalization. Augmenting training data with additional information from other counties significantly improves performance, with neighbor-based and same-type-based data augmentation being effective. However, as we trained these models without explicitly distinguishing between data from different counties, the current approach still leaves room for improvement. This calls for more sophisticated methods to better utilize and integrate information from various counties while maintaining awareness of their unique temporal and spatial characteristics.

### 2.2 Differences in County Characteristics and Predictive Challenges

In this subsection, we explore how model performance and prediction difficulty vary across counties, particularly based on population size. By distinguishing between large and small counties, we aim to understand the factors driving disparities in predictive accuracy and how these differences can guide the design of effective data augmentation and model training strategies.

To illustrate the stark contrast in overdose death trends, we present the quarterly opioid overdose death counts for Franklin County, a large urban county, and Harrison County, a smaller rural county, in Figure 1. Figure 1 reveals a clear divergence in magnitude: Franklin County consistently records over 100 deaths per quarter, whereas Harrison County typically records fewer than 3 deaths per quarter. A horizontal line at *y* = 3 highlights this threshold, demonstrating that small counties frequently hover near or below this critical point. This observation underscores the inherent challenge with small counties—predicting minor variations in overdose deaths becomes highly sensitive and volatile. For instance, in Harrison County, a shift from 2 to 3 deaths per quarter represents a 50% change, a significant fluctuation under regression metrics like RMSE. Conversely, in Franklin County, a similar absolute change (from 40 to 41 deaths) is negligible and barely impacts the overall error. This difference highlights how small counties inherently exhibit noisier and more volatile trends, making them much harder to predict with relative continuous patterns.

**Figure 1.**
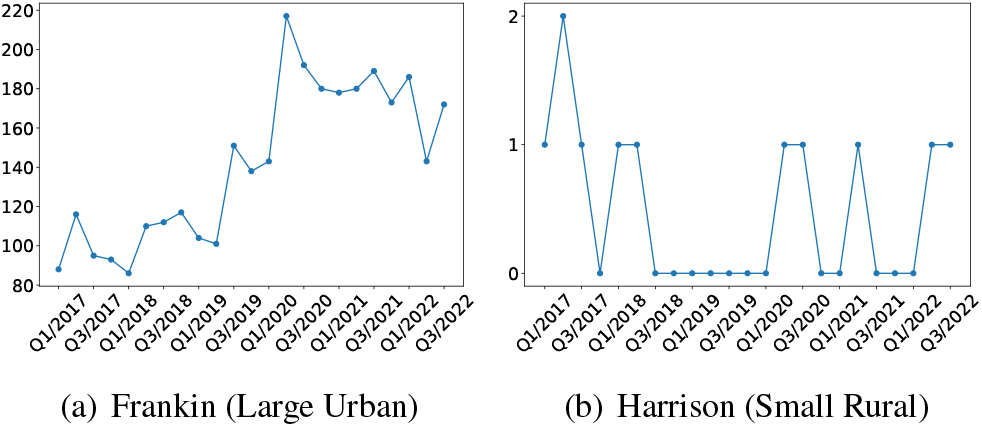
Quarterly death count comparison.

To further investigate how county size affects model performance, we grouped counties into four types: Appalachian (smallest), rural, suburban, and urban (largest), and evaluated the LSTM performance on each category. Table 2 compares the RMSE of the LSTM across these county types. The model achieves the lowest RMSE on urban and suburban counties, reflecting their more predictable and stable trends. In contrast, performance degrades significantly in rural and Appalachian counties, where smaller populations and lower death counts introduce greater uncertainty. This performance gap stems from the inherent difficulty of making accurate predictions in small counties, where death counts often hover near zero or just a few cases per quarter. As previously noted, even a small prediction error in these counties can lead to large relative percentage errors due to the small denominator—a limitation of using RMSE for regression tasks involving low-count events. Moreover, applying data augmentation uniformly across counties can exacerbate this issue. While data augmentation may improve performance for small counties, it risks causing performance drops in large counties due to the potential introduction of noise from poorly generalized patterns.

**Table 2.** Performance comparison of four county types using LSTM.

|  | Appalachian |  | Rural |  | Suburban |  | Urban |  | All |  |
| --- | --- | --- | --- | --- | --- | --- | --- | --- | --- | --- |
|  | RMSE | SMAPE | RMSE | SMAPE | RMSE | SMAPE | RMSE | SMAPE | RMSE | SMAPE |
| LSTM | 9.731 | 0.786 | 17.300 | 0.740 | 5.321 | 0.625 | 6.071 | 0.306 | 11.987 | 0.674 |

The challenge posed by small counties is further illustrated in Figure 2, which presents the population distributions of Appalachian (smallest) and urban (largest) counties, respectively. The bar charts reveal a stark contrast in the number of individuals residing in these regions. Urban counties are densely populated, offering a larger and more stable data sample for the model to learn from, whereas Appalachian counties have sparse populations, making them susceptible to data sparsity and instability. This imbalance exacerbates prediction difficulties, as limited data in small counties often lacks the temporal and spatial diversity needed for effective generalization.

**Figure 2.**
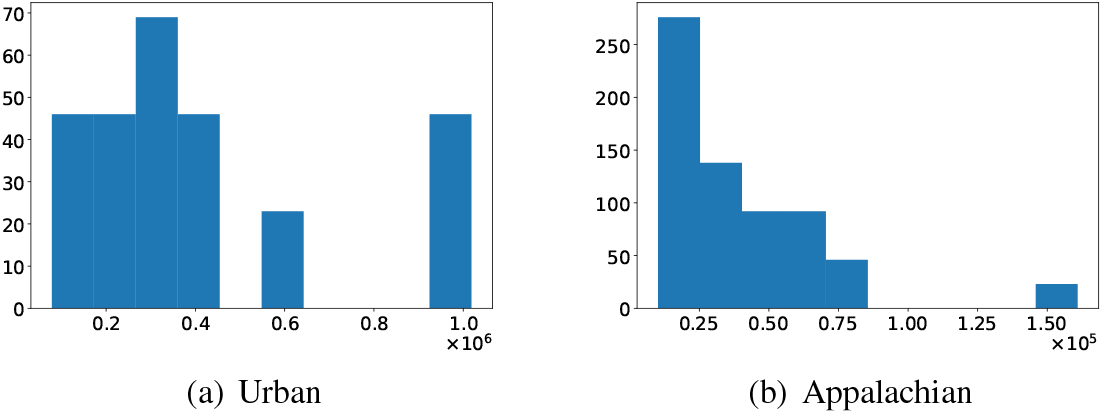
Population distribution of Appalachian and Urban counties.

#### Key Takeaway

Given these observations, it is clear that a one-size-fits-all approach to data augmentation and model training is insufficient. Small counties, such as those in appalachian and rural regions, require specialized treatment due to their high volatility and sensitivity to prediction errors. For instance, small counties can benefit from a binary classification task that predicts whether the death count will exceed a threshold (e.g., 3 cases) rather than directly regressing on precise death counts, which are prone to large errors. For larger counties, like those in urban and suburban areas with abundant and stable data, traditional regression tasks are more suitable. Moreover, to address the challenges of data augmentation, it is important to design strategies that inform the model about the origin of the augmented data. Small counties should be able to leverage data from large counties to improve their generalization ability, but exposure of large counties to data from small counties should be limited to avoid introducing noise. This targeted approach to data augmentation ensures that small counties can benefit from the diversity and stability of large-county data, while large counties remain protected from the volatility of small-county data. By leveraging this differentiated strategy, we can enhance robustness and accuracy across counties of varying sizes and characteristics.

## 3 Problem Formulation

### 3.1 System Setup

We define a graph-based temporal system to predict county-level OD death rates by modeling both spatial relationships and temporal dynamics. Let *G*_*t*_ = *{V, E, X*_*t*_*}* denote a dynamic graph at time *t*, where:

1. *V* = *{v*_1_, *v*_2_, …, *v*_|*V* |_*}* represents the set of nodes, with each node *v*_*i*_ corresponding to an Ohio county.
2. *E* denotes the set of edges representing adjacent county connections derived from geographic proximity.
3. *X*_*t*_ denotes the node feature matrix at time *t*, where each node *v*_*i*_ is associated with a feature vector *x*_*i,t*_ that comprises both dynamic and static attributes, as summarized in Table 6.

The goal is to predict the future opioid overdose death rate *y*_*i,t*+1_ for each county *v*_*i*_ by leveraging historical information from *X*_*t*_, as well as the hidden representations propagated through spatial neighbors.

### 3.2 Problem Objective

Let 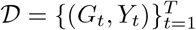 denote the time-series dataset, where *Y*_*t*_ represents the observed overdose death rates at time *t*.

The problem is formulated as learning a spatio-temporal graph neural network *f*_*θ*_ parameterized by *θ* to predict future overdose death rates by minimizing the following objective function:

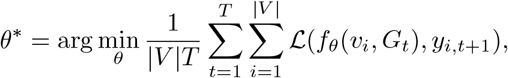

where ℒ (·) is a task-specific loss function, such as Mean Squared Error (MSE) for regression tasks or Binary Cross-Entropy Loss (BCE) for classification tasks.

## 4. Methodology

Our proposed Spatial-Temporal Graph Neural Network (ST-GNN) framework integrates temporal sequence modeling and graph-based spatial learning to predict county-level opioid overdose deaths. As illustrated in Figure 3, the architecture consists of three main components: temporal embedding generation, spatial message aggregation, and final prediction output.

**Figure 3.**
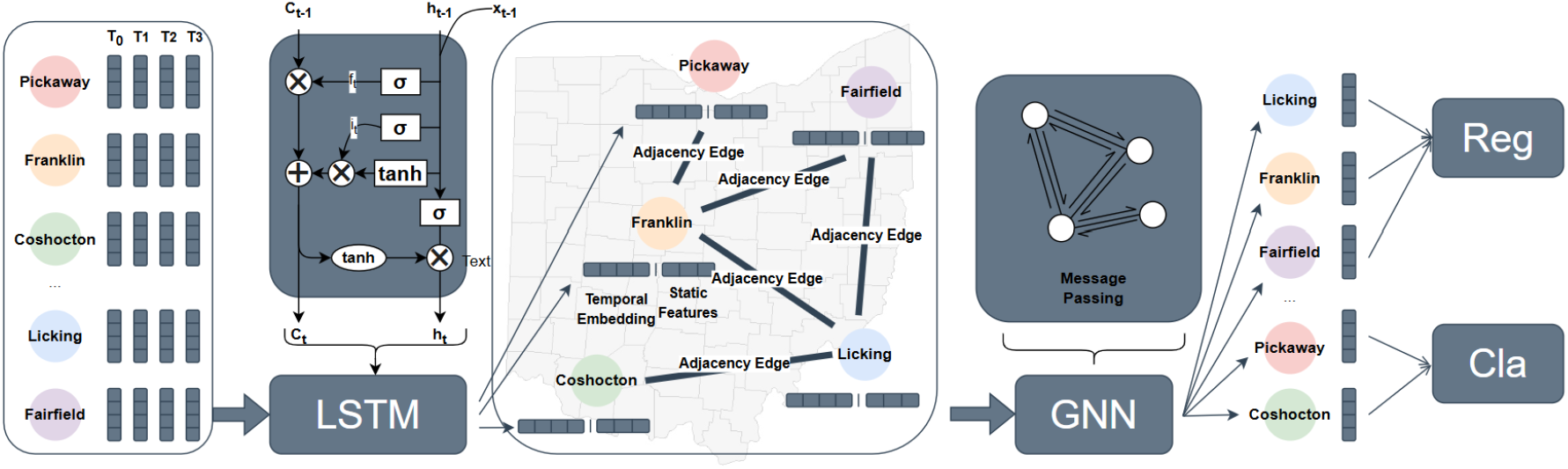
Illustration of the proposed method. Dynamic temporal features are first processed through an LSTM to generate temporal embeddings for each node. The elementwise operations are shown in circles, while the non-linear layers of the LSTM are depicted in squares. The generated embeddings are then concatenated with static features to form the initial node embeddings, which are passed through the GNN for message aggregation. This process generates the hidden embeddings for each county, which are then utilized to produce the final regression output (Reg) for large counties like Franklin, or classification (Cla) for small counties like Coshocton.

### Temporal Feature Processing with LSTM

To capture the historical patterns of opioid overdose deaths and related risk factors, we first process the dynamic temporal features through a Long Short-Term Memory (LSTM) network. Each county’s time-series input consists of quarterly opioid-related indicators, including naloxone administration events, high-risk opioid prescribing, and other epidemiological signals. The LSTM generates a temporal embedding for each county by extracting sequential dependencies from its past observations.

### Feature Fusion and Node Representation

The temporal embeddings from the LSTM are concatenated with static county-level features, which corresponds to the SDoH index. This fusion forms a comprehensive node representation that encodes both long-term socioeconomic conditions and short-term opioid-related fluctuations. The resulting vector serves as the initial node embedding for the subsequent spatial step.

### Graph Neural Network for Spatial Dependency Modeling

The generated node embeddings are passed through a Graph Neural Network (GNN) to model spatial dependencies between counties. The adjacency graph, constructed from geographic proximity, enables counties to exchange information with their neighbors. The GNN aggregates features from connected nodes using message-passing mechanisms, allowing each county’s representation to be enriched with spatially relevant data. Our proposed framework is compatible with multiple GNN variants, including Graph Attention Networks (GAT) [21] and Graph Convolutional Networks (GCN) [22].

### Dual Training Loss

To account for population size disparities among counties, we adopt a dual-task learning approach: For large counties (e.g., Franklin County), where opioid overdose death counts are relatively high and stable, the model performs regression, predicting the exact number of deaths in the next quarter. For small counties (e.g., Coshocton County), where overdose deaths are sparse and volatile, we frame the task as binary classification, predicting whether the count will exceed a threshold (e.g., three deaths in a quarter).

Specifically, the loss function is formulated as follows:

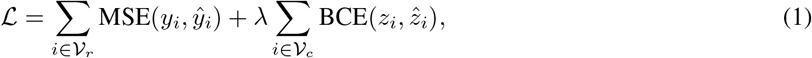

where *y*_*i*_ is the ground truth overdose death count for county *i, ŷ*_*i*_ is the predicted overdose death count for county *i. z*_*i*_ is the binary label for small counties, defined as:

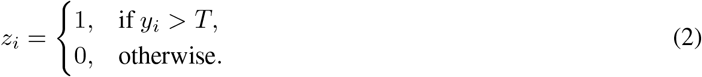

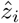 is the predicted probability of exceeding *T* deaths for small counties. In this work, we set *T* = 3. And *λ* is the hyperparameter that controls the balance between regression and classification tasks.

This joint loss function ensures that the model adapts to counties with different data distributions, mitigating the issue of overfitting in low-count regions while maintaining precision in high-count areas.

## 5 Experiment and Results

### 5.1 Experimental Setting

We applied our proposed Spatial-Temporal Graph Neural Network (ST-GNN) framework to predict the opioid OD death rate for each of Ohio’s 88 counties using quarterly data spanning from Q1 2017 to Q2 2023. The OD rate for each county was standardized by dividing the death count by the population per 100,000, ensuring comparability across counties of varying sizes. The dataset was split into three temporal segments: data from Q1 2017 to Q3 2020 was used for model training, data from Q4 2020 to Q4 2021 for validation, and data from Q1 2022 onwards for testing.

The time window for historical data was set to four consecutive quarters, capturing seasonality and temporal dependencies crucial for understanding the evolution of opioid-related events. Our experiments utilized a graph comprising 88 nodes (representing counties) and 238 edges, where edges were defined based on geographic adjacency between counties. This setup allowed the model to capture spatial dependencies through message passing between adjacent counties.

The model was implemented using Python with PyTorch Geometric (PyG) version 2.5.2, leveraging its efficient graph processing capabilities. We evaluated the performance using two key metrics: Root Mean Square Error (RMSE) and

Symmetric Mean Absolute Percentage Error (SMAPE). RMSE measures the absolute magnitude of prediction errors, while SMAPE provides a percentage-based assessment, making it particularly suitable for datasets with both high and low OD rates.

To assess the effectiveness of our proposed framework, we compared its performance against several baseline models, including LSTM[20], DCRNN [23] and GConvLSTM[24], which are well-established temporal graph neural network models designed for time-series forecasting in spatial contexts. This comparative evaluation highlights the strengths of ST-GNN in capturing both temporal and spatial dependencies for predicting opioid overdose deaths.

### 5.2 Results

As shown in Table 3, the proposed ST-GNN consistently outperforms both traditional statistical models (ARIMA) and deep learning baselines (LSTM, DCRNN, and GConvLSTM). For large counties, performance is evaluated using RMSE and SMAPE, where RMSE measures absolute prediction error magnitude and SMAPE captures relative error scaled by observed counts. ST-GAT achieves the lowest RMSE, indicating improved accuracy in predicting continuous death rates, while the edge-weighted ST-GCN attains the best SMAPE, reflecting better relative error control. For small counties, we evaluate classification performance using ROC-AUC and F1 score, where ROC-AUC measures discrimination ability across thresholds and F1 balances precision and recall under class imbalance. The edge-weighted ST-GCN achieves the highest ROC-AUC, while ST-GAT attains the best F1 score, demonstrating improved robustness in sparse and volatile settings. These results confirm that jointly modeling spatial and temporal dependencies, together with our dual-task loss formulation, improves predictive stability across heterogeneous county populations.

**Table 3.** Performance comparison of different models. ST-GCN* represents ST-GCN model trained with edge weights. The best performance is highlighted.

|  |  | Baselines |  |  |  | Ours |  |  |
| --- | --- | --- | --- | --- | --- | --- | --- | --- |
|  |  | ARIMA | LSTM | DCRNN | GConvLSTM | ST-GCN | ST-GCN* | ST-GAT |
| Large Counties | RMSE (↓) | 18.322 | 12.998 | 14.519 | 15.866 | 9.253 | 9.348 | **9.149** |
| | SMAPE (↓) | 0.583 | 0.362 | 0.404 | 0.437 | 0.256 | **0.242** | 0.244 |
| Small Counties | ROC-AUC (↑) | 0.508 | 0.521 | 0.502 | 0.511 | 0.702 | **0.738** | 0.714 |
| | F1 (↑) | 0.452 | 0.483 | 0.518 | 0.521 | 0.552 | 0.554 | **0.579** |

### 5.3 Discussion

#### Ablation Study on Loss Function

To evaluate the effectiveness of our dual-task loss function, we conduct an ablation study by separately training our models with only regression loss (Reg) for all counties, only classification loss (Cla) for small counties, and our full combined loss (Both). We present the results in Table 4.

**Table 4.** Ablation study of loss type. “Reg” denotes models trained solely with regression loss, while “Cla” represents models trained only with classification loss. The regression evaluation metrics for “Cla” models on large counties are omitted, as they are not meaningful. For small counties, we derive classification performance from “Reg” models by treating regressed values greater than 3 as a classification of 1 and values of 3 or below as 0.

|  |  | ST-GCN |  |  | ST-GAT |  |  |
| --- | --- | --- | --- | --- | --- | --- | --- |
|  |  | Reg | Cla | Both | Reg | Cla | Both |
| Large Counties | RMSE (↓) | 11.001 | - | 9.253 | 11.572 | - | **9.149** |
| | SMAPE (↓) | 0.486 | - | 0.256 | 0.518 | - | **0.245** |
| Small Counties | ROC-AUC(↑) | 0.601 | 0.446 | 0.702 | 0.555 | 0.461 | **0.714** |
| | F1 (↑) | 0.539 | 0.496 | 0.552 | 0.532 | 0.527 | **0.579** |

Models trained with only regression loss (Reg) perform significantly worse in large counties, as evidenced by higher RMSE compared to the dual-task models. Likewise, using only classification loss (Cla) degrades performance for small counties, resulting in lower ROC-AUC and F1 scores. In contrast, the combined loss function (Both) consistently achieves the best performance, demonstrating its effectiveness in balancing the heterogeneous prediction tasks across counties of different sizes.

Without explicitly distinguishing between county types, the two tasks interfere with each other, leading to suboptimal performance for both large and small counties. More specifically, regression models (Reg) struggle in small counties, as they lack the capacity to handle low and volatile overdose counts, making predictions unstable. Classification models (Cla) fail in large counties, as they reduce the problem to a binary decision, losing the ability to capture fine-grained variations in overdose rates, which are essential for public health interventions. The joint loss model (Both) effectively resolves these issues by allowing large counties to benefit from precise regression modeling, capturing subtle trends in overdose deaths. It also allows small counties to leverage classification robustness, improving stability in cases where overdose counts fluctuate near zero or remain sparse. This targeted approach ensures that the model adapts appropriately to counties of different sizes, maximizing overall prediction accuracy.

#### Hyper-parameter Study

To evaluate the impact of *λ*, which balances regression for large counties and classification for small counties, we tested values 10, 30, 50, and 100. As shown in Table 5, *λ* = 30 achieves the best overall performance. A smaller *λ* yields slightly better RMSE but struggles with small-county volatility. In contrast, a larger *λ* results in higher F1 but worsens RMSE and SMAPE, as over-prioritizing small-county signals reduces regression accuracy. Thus, we conclude that *λ* = 30 provides the best balance, ensuring that neither task dominates the other.

**Table 5.**
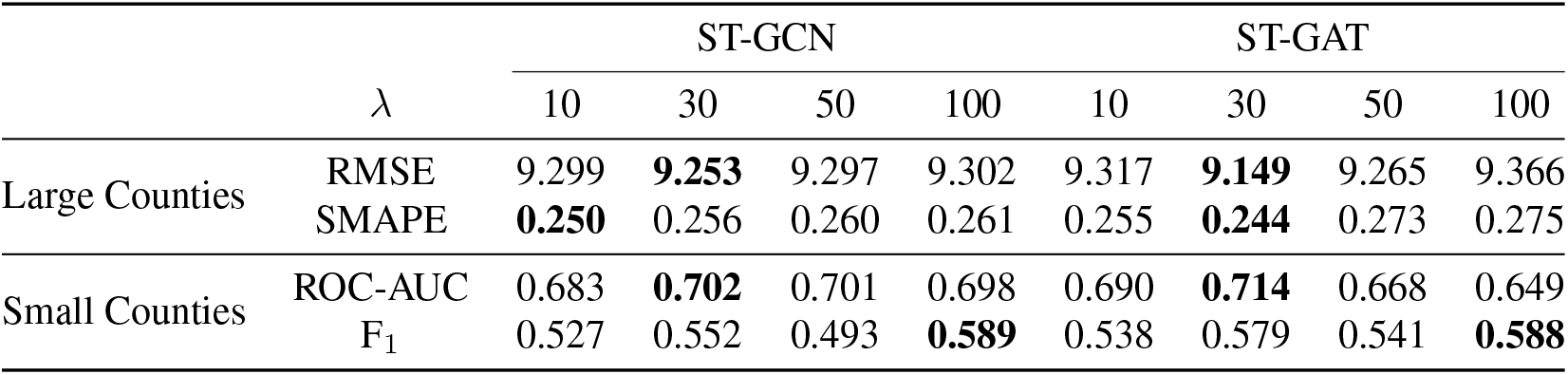
Hyper-parameter study of *λ*.

**Table 6.** Features used for opioid OD death prediction.

| Category | Feature |
| --- | --- |
| Dynamic Feature | Drug OD deaths |
| Dynamic Feature | EMS events involving naloxone administration |
| Dynamic Feature | EMS events with suspected opioid poisoning |
| Dynamic Feature | Incident high-risk opioid prescribing |
| Dynamic Feature | Individuals receiving buprenorphine as MOUD/MAT |
| Dynamic Feature | Individuals receiving buprenorphine products for 6+ months |
| Dynamic Feature | Naloxone units distributed by Project DAWN in county |
| Dynamic Feature | New acute opioid prescriptions limited to 7-day supply |
| Dynamic Feature | New continual opioid treatment lasting 31+ days |
| Static Feature | SDI score |
| Static Feature | Percentage of poverty below 100% Federal Poverty Level |
| Static Feature | Percentage of single-parent families |
| Static Feature | Percentage of low education attainment (adults with less than 12 years of education) |
| Static Feature | Percentage of households without a vehicle |
| Static Feature | Percentage of renter-occupied households (rather than owner-occupied) |
| Static Feature | Percentage of household crowding (more than one person per room) |
| Static Feature | Percentage of non-employed individuals |

## 6. Conclusion

In this study, we proposed a Spatial-Temporal Graph Neural Network (ST-GNN) framework to predict opioid overdose death rates at the county level in Ohio. By integrating GNNs for spatial relationships and LSTM for temporal dynamics, our model effectively captures spatio-temporal dependencies, outperforming traditional time-series and naive graph-based methods. Our results demonstrate that joint modeling of spatial and temporal features significantly enhances prediction accuracy, particularly when incorporating edge-weighted GNNs or attention-based aggregation. Additionally, we introduced a dual-task loss function, applying regression for large counties and classification for small counties, which improves performance across counties of varying sizes. Overall, this work highlights the importance of tailored spatio-temporal modeling in addressing the opioid crisis. Future research could explore incorporating external risk factors, enhancing model interpretability, and extending the framework to other regions. By refining predictive models for opioid overdose deaths, we aim to support data-driven public health interventions and policy decisions.

## Data Availability

All data used in this study are publicly available. Overdose mortality and related indicators are available from the Ohio Integrated Behavioral Health Dashboard maintained by the Ohio Department of Health (https://data.ohio.gov/wps/portal/gov/data/view/ohio-ibhd). Social determinants of health variables were obtained from the U.S. Census Bureau American Community Survey (https://www.census.gov/programs-surveys/acs).

https://www.census.gov/programs-surveys/acs

## Acknowledgement

We acknowledge the funding for the project provided by the National Institute on Drug Abuse (R01DA057668) and the National Science Foundation (2145625).

## A Feature Description

## References

1. Jonathan Penm, Neil J MacKinnon, Jill M Boone, Antonio Ciaccia, Cameron McNamee, and Erin L Winstanley. Strategies and policies to address the opioid epidemic: a case study of ohio. Journal of the American Pharmacists Association, 57(2):S148–S153, 2017.

2. Chihyun Park, Jean R Clemenceau, Anna Seballos, Sara Crawford, Rocio Lopez, Tyler Coy, Gowtham Atluri, and Tae Hyun Hwang. A spatiotemporal analysis of opioid poisoning mortality in ohio from 2010 to 2016. Scientific reports, 11(1):4692, 2021.

3. Vaishali S Deo, Manreet K Bhullar, Thomas P Gilson, Daniel J Flannery, and Sarah E Fulton. The need to rethink harm reduction for people using drugs alone to reduce overdose fatalities. Substance Use & Misuse, 59 (3):450–458, 2024.

4. Andres Hernandez, Adam J Branscum, Jingjing Li, Neil J MacKinnon, Ana L Hincapie, and Diego F Cuadros. Epidemiological and geospatial profile of the prescription opioid crisis in ohio, united states. Scientific reports, 10(1):4341, 2020.

5. Ohio Department of Health. 2022 unintentional drug overdose annual report, 2023. URL https://shorturl.at/RsDD0.

6. Jon E Zibbell, Arnie P Aldridge, Dennis Cauchon, Jolene DeFiore-Hyrmer, and Kevin P Conway. Association of law enforcement seizures of heroin, fentanyl, and carfentanil with opioid overdose deaths in ohio, 2014-2017. JAMA Network Open, 2(11):e1914666–e1914666, 2019.

7. Christine A Schalkoff, Kathryn E Lancaster, Bradley N Gaynes, Vivian Wang, Brian W Pence, William C Miller, and Vivian F Go. The opioid and related drug epidemics in rural appalachia: A systematic review of populations affected, risk factors, and infectious diseases. Substance abuse, 41(1):35–69, 2020.

8. Steven A Sumner, Daniel Bowen, Kristin Holland, Marissa L Zwald, Alana Vivolo-Kantor, Gery P Guy, William J Heuett, DeMia P Pressley, and Christopher M Jones. Estimating weekly national opioid overdose deaths in near real time using multiple proxy data sources. JAMA network open, 5(7):e2223033–e2223033, 2022.

9. Lindsey M Ferris, Brendan Saloner, Noa Krawczyk, Kristin E Schneider, Molly P Jarman, Kate Jackson, B Casey Lyons, Matthew D Eisenberg, Tom M Richards, Klaus W Lemke, et al. Predicting opioid overdose deaths using prescription drug monitoring program data. American journal of preventive medicine, 57(6):e211–e217, 2019.

10. Wei-Hsuan Lo-Ciganic, James L Huang, Hao H Zhang, Jeremy C Weiss, Yonghui Wu, C Kent Kwoh, Julie M Donohue, Gerald Cochran, Adam J Gordon, Daniel C Malone, et al. Evaluation of machine-learning algorithms for predicting opioid overdose risk among medicare beneficiaries with opioid prescriptions. JAMA network open, 2(3):e190968–e190968, 2019.

11. George Panagopoulos, Giannis Nikolentzos, and Michalis Vazirgiannis. Transfer graph neural networks for pandemic forecasting. In Proceedings of the AAAI Conference on Artificial Intelligence, volume 35, pages 4838–4845, 2021.

12. Lijing Wang, Aniruddha Adiga, Jiangzhuo Chen, Adam Sadilek, Srinivasan Venkatramanan, and Madhav Marathe. Causalgnn: Causal-based graph neural networks for spatio-temporal epidemic forecasting. In Proceedings of the AAAI conference on artificial intelligence, volume 36, pages 12191–12199, 2022.

13. Cornelius Fritz, Emilio Dorigatti, and David Rügamer. Combining graph neural networks and spatio-temporal disease models to improve the prediction of weekly covid-19 cases in germany. Scientific Reports, 12(1):3930, 2022.

14. Avijit Mitra, Hiba Ahsan, Wenjun Li, Weisong Liu, Robert D Kerns, Jack Tsai, William Becker, David A Smelson, Hong Yu, et al. Risk factors associated with nonfatal opioid overdose leading to intensive care unit admission: a cross-sectional study. JMIR medical informatics, 9(11):e32851, 2021.

15. Nina Cesare, Lisa M Lines, Redonna Chandler, Erin B Gibson, Rachel Vickers-Smith, Rebecca Jackson, Angela R Bazzi, Dawn Goddard-Eckrich, Nasim Sabounchi, Deena J Chisolm, et al. Development and validation of a community-level social determinants of health index for drug overdose deaths in the healing communities study. Journal of Substance Use and Addiction Treatment, 157:209186, 2024.

16. Jason M Glanz, Komal J Narwaney, Shane R Mueller, Edward M Gardner, Susan L Calcaterra, Stanley Xu, Kristin Breslin, and Ingrid A Binswanger. Prediction model for two-year risk of opioid overdose among patients prescribed chronic opioid therapy. Journal of general internal medicine, 33:1646–1653, 2018.

17. Xinyu Dong, Jianyuan Deng, Wei Hou, Sina Rashidian, Richard N Rosenthal, Mary Saltz, Joel H Saltz, and Fusheng Wang. Predicting opioid overdose risk of patients with opioid prescriptions using electronic health records based on temporal deep learning. Journal of biomedical informatics, 116:103725, 2021.

18. Wei-Hsuan Lo-Ciganic, Julie M Donohue, Qingnan Yang, James L Huang, Ching-Yuan Chang, Jeremy C Weiss, Jingchuan Guo, Hao H Zhang, Gerald Cochran, Adam J Gordon, et al. Developing and validating a machine-learning algorithm to predict opioid overdose in medicaid beneficiaries in two us states: a prognostic modelling study. The Lancet Digital Health, 4(6):e455–e465, 2022.

19. F. B. Ahmad, J. A. Cisewski, L. M. Rossen, and P. Sutton. Provisional drug overdose death counts, 2025. URL 10.15620/cdc/20250305008.

20. Sepp Hochreiter and Jürgen Schmidhuber. Long short-term memory. Neural Comput., 1997. ISSN 0899-7667. doi: 10.1162/neco.1997.9.8.1735. URL https://doi.org/10.1162/neco.1997.9.8.1735.

21. Petar Veličković, Guillem Cucurull, Arantxa Casanova, Adriana Romero, Pietro Liós, and Yoshua Bengio. Graph Attention Networks. ICLR, 2018.

22. Thomas N Kipf and Max Welling. Semi-supervised classification with graph convolutional networks. ICLR, 2017.

23. Yaguang Li, Rose Yu, Cyrus Shahabi, and Yan Liu. Diffusion convolutional recurrent neural network: Data-driven traffic forecasting, 2018.

24. Youngjoo Seo, Michaël Defferrard, Pierre Vandergheynst, and Xavier Bresson. Structured sequence modeling with graph convolutional recurrent networks, 2017.

